# Genetically Predicted Blood DNA Methylation Reveals Putative Regulatory Signals Associated with ALS Risk

**DOI:** 10.64898/2026.09.17.26363319

**Authors:** Tianying Zhao, Mariah Hoffman, Gang Wu, Veronique Belzil

**Author notes:** **Correspondence to**: Veronique Belzil, MS, PhD, Associate Professor of Neurology & Genetic Medicine, Director, Vanderbilt ALS Research Center, Vanderbilt University Medical Center. **Email addresses:** Tianying Zhao, Mariah Hoffman, Gang Wu, Veronique Belzil.

## Abstract

Amyotrophic lateral sclerosis (ALS) is a fatal neurodegenerative disorder whose genetic architecture and underlying molecular mechanisms remain incompletely understood, particularly in sporadic disease. To investigate whether genetically regulated DNA methylation may help interpret ALS susceptibility, we conducted a methylome-wide association study (MWAS) of genetically predicted blood DNA methylation using the PrediXcan framework. CpG-specific prediction models developed in the ARIES and Understanding Society cohorts were applied to ALS genome-wide association study (GWAS) summary statistics from 27,205 cases and 110,881 controls of European ancestry. In total, genetically predicted methylation at 192,378 unique CpG sites was evaluated using S-PrediXcan. At a nominal threshold of p < 0.05, 3,741 CpGs were associated with ALS risk using ARIES models and 13,127 using Understanding Society models. After Bonferroni correction, 25 CpGs across eight genomic regions remained significantly associated with ALS risk. These included signals near established ALS and ALS-frontotemporal dementia genes and loci, including *C9orf72*, *TBK1*, *SCFD1*, and *MOB3B*, as well as three CpGs mapping to *WHAMM* at 15q25.2, a region not previously implicated in ALS by GWAS. Predicted methylation was positively associated with ALS risk at 18 CpGs and inversely associated at seven. Complementary transcriptome-wide association analyses using GTEx v8 whole-blood gene-expression prediction models identified 11 genes associated with ALS risk after Bonferroni correction, including convergent methylation and expression signals at *C9orf72*. These findings add a regulatory dimension to ALS genetic studies by prioritizing CpG sites, genes, and genomic regions through which inherited variation may influence disease susceptibility. PrediXcan-based MWAS therefore provides a complementary strategy for refining genetic association signals into biologically testable candidates and identifying regulatory mechanisms for further functional investigation.

## Introduction

Amyotrophic lateral sclerosis (ALS) is a fatal neurodegenerative disorder characterized by progressive motor neuron loss, muscle weakness, paralysis, and respiratory failure, with a median survival of approximately 2.5 years from symptom onset [1]. Diagnosis remains challenging because early symptoms often overlap with those of other conditions [2], and patients may not encounter a neurologist until relatively late in the diagnostic journey [3,4].

ALS is clinically heterogeneous, with upper and lower motor neuron dysfunction affecting bulbar, cervical, thoracic, or lumbar regions and progressively impairing limb movement, swallowing, speech, and respiratory function [2,5]. Cognitive and behavioral changes may also arise early in the disease course [6,7] and are recognized in approximately 35–50% of individuals with ALS [8,9]. This combination of rapid progression and substantial clinical heterogeneity underscores the need to better understand the biological mechanisms that influence ALS susceptibility and disease expression.

Approximately 10% of ALS cases are classified as familial, whereas the remaining 90% are considered sporadic [10]. An underlying Mendelian mutation can be identified in approximately 50–85% of familial ALS cases [11] and in up to 10–20% of apparently sporadic cases [12–14]. Thus, although known pathogenic variants explain a substantial proportion of familial disease, the genetic architecture underlying ALS susceptibility remains incompletely understood, particularly among sporadic cases.

Large-scale genetic studies have substantially advanced the identification of ALS susceptibility loci. The largest ALS genome-wide association study (GWAS) to date included 29,612 individuals with ALS and 122,656 controls and identified multiple loci associated with disease risk [15]. However, GWAS signals alone often provide limited information about the molecular mechanisms through which associated variants influence disease susceptibility, particularly because many risk variants occur in noncoding regions. Integrative approaches that connect inherited genetic variation with intermediate molecular traits may therefore help translate statistical associations into biologically interpretable signals [16].

PrediXcan was developed to address this challenge by estimating genetically regulated gene expression from genotype data using prediction models trained in reference transcriptomic datasets and subsequently testing predicted expression for association with phenotypes [16]. S-PrediXcan extends this framework to GWAS summary statistics, allowing gene-level associations to be estimated without access to individual-level genotype and phenotype data [17]. PrediXcan and S-PrediXcan have shown high concordance, supporting the use of summary-statistic approaches for large-scale molecular association studies [17].

Although initially developed for transcriptome-wide association studies (TWAS), the same framework can be extended to other genetically regulated molecular traits. DNA methylation at many CpG sites is influenced by inherited genetic variation [18–21], exhibits a significant single-nucleotide polymorphism (SNP)-based heritable component [21–23], and is associated with specific genetic variants through methylation quantitative trait loci [19,24–26]. These properties make a genetically regulated component of DNA methylation predictable from genotype.

Building on this concept, Fryett et al. adapted the PrediXcan framework to model genetically predicted DNA methylation and test its association with complex traits [27]. Using matched genotype and methylation data from the Accessible Resource for Integrated Epigenomics Studies (ARIES) [28] and the Understanding Society study [29], they developed prediction models for CpG sites whose methylation levels could be reliably estimated from nearby genetic variation [27]. This approach provides an opportunity to investigate whether the genetically regulated component of DNA methylation is associated with complex disease risk without requiring methylation measurements in the GWAS population itself.

Here, we conducted a methylome-wide association study (MWAS) by applying these established ARIES and Understanding Society methylation prediction models to ALS GWAS summary statistics from 27,205 cases and 110,881 controls of European ancestry [15] using S-PrediXcan [17]. We tested whether genetically predicted blood DNA methylation at individual CpG sites was associated with ALS susceptibility and complemented these analyses with TWAS using GTEx v8 whole-blood gene-expression prediction models [30]. By integrating ALS genetic association data with genetically regulated methylation and gene expression, our goal was not to identify diagnostic biomarkers or establish causal methylation changes, but to prioritize CpG sites, genes, and genomic regions that may help explain how inherited genetic variation influences ALS risk. This strategy adds a regulatory layer to conventional GWAS interpretation and may help refine broad genetic association signals into biologically testable candidates for functional investigation.

## Materials and Methods

### Study population

We used ALS GWAS summary statistics from 27,205 cases and 110,881 controls of European ancestry [15], representing the largest ALS GWAS dataset available to date. The GWAS combined individual-level genotype data from 117 cohorts into six strata matched by genotyping platform, with standard quality-control procedures applied prior to association analysis [15].

### DNA methylation prediction models

We used previously developed CpG-specific DNA methylation prediction models from Fryett et al. [27] for association analyses with ALS risk (Figure 1). Model development has been described in detail previously [27]. Briefly, the models were trained using samples with matched genotype and DNA methylation data from the ARIES [28] and Understanding Society [29] cohorts. Following quality control, 841 ARIES mother samples and 1,120 Understanding Society samples were available for model development.

**Figure 1.**
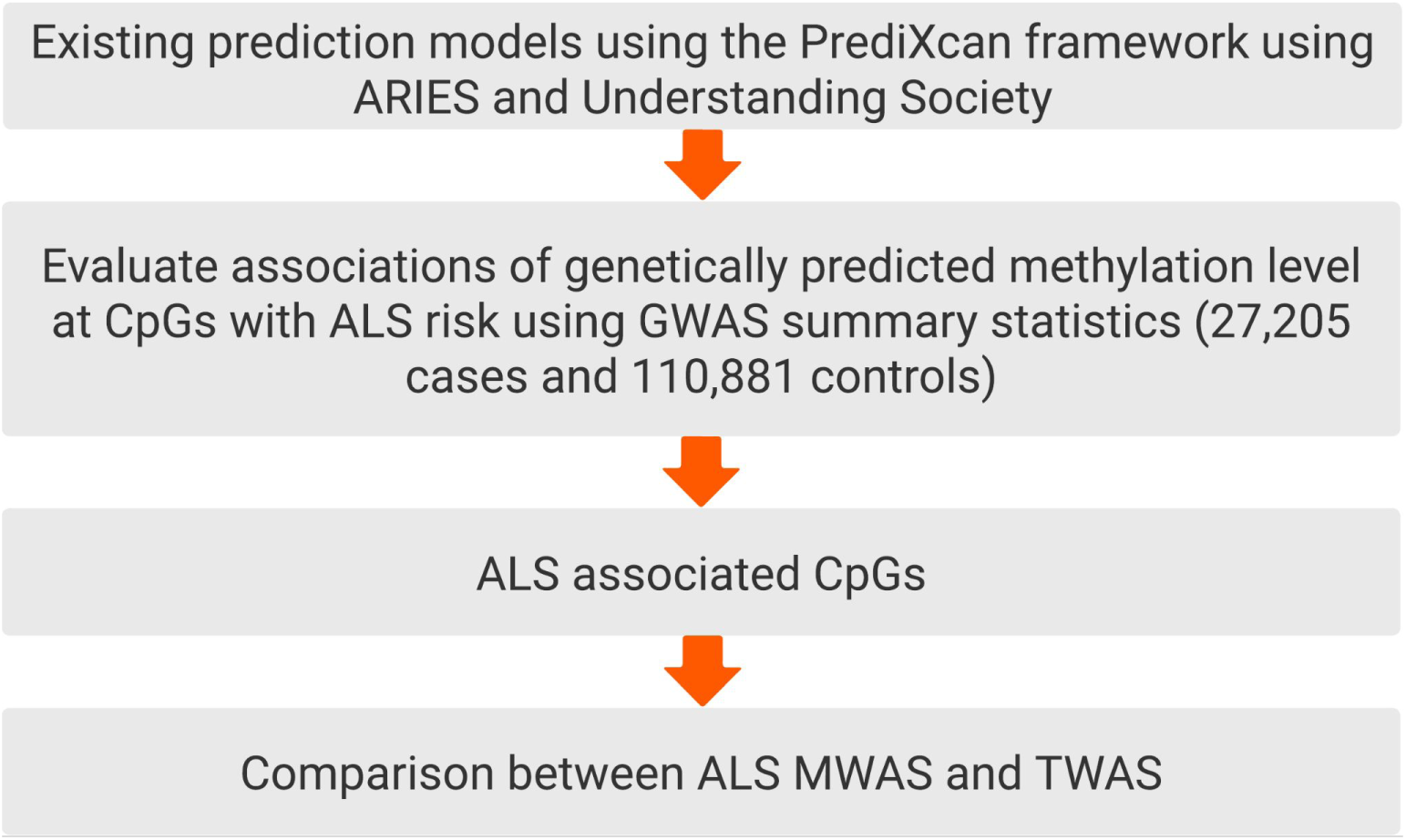
Study design flowchart.

Three penalized regression approaches were evaluated: ridge regression [31], LASSO [32], and elastic net [33] with α = 0.5. For each approach, the regularization parameter λ was selected using 10-fold cross-validation to minimize the mean squared error between predicted and observed methylation among models containing at least one SNP. Prediction performance was assessed using the correlation (R) between predicted and observed methylation levels, and elastic net with α = 0.5 was selected as the optimal modeling approach.

Five cis-window sizes surrounding each CpG (250 kb, 500 kb, 1 Mb, 2 Mb, and 3 Mb) were subsequently evaluated, with the optimal window selected separately for each CpG site. Final models were trained using all available samples within each cohort and externally validated in the other cohort. Only models achieving a validation correlation of R ≥ 0.1 were retained for downstream analyses.

### Gene expression prediction models

We used GTEx v8 elastic net transcriptome prediction models developed by Barbeira et al. [16,17,30]. These models were trained using matched GTEx v8 RNA-seq and genotype data from individuals of European ancestry and were restricted to genes annotated in GENCODE v26 as protein-coding, long noncoding RNA, or pseudogenes [34].

For each gene–tissue pair, adjusted gene expression was modeled as a function of nearby genetic variation using elastic net regression. Candidate variants were restricted to HapMap 3 CEU SNPs with minor allele frequency > 0.01 located within 1 Mb upstream of the transcription start site or 1 Mb downstream of the transcription end site. Models were fitted using *glmnet* [35] with α = 0.5, and the penalty parameter was selected by 10-fold cross-validation. Models were retained when the mean Pearson correlation between predicted and observed expression across the 10 folds exceeded 0.1 and the nested cross-validated correlation test reached p < 0.05.

### Association analysis of genetically predicted DNA methylation levels with ALS risk

DNA methylation prediction models were included in downstream association analyses if they met predefined performance criteria of R > 0.1 and p < 0.05, consistent with thresholds used in a previously published study [36]. Each retained CpG-specific prediction model was applied to the ALS GWAS summary statistics [15] using S-PrediXcan [17], following the previously described implementation [37]. For each CpG site, S-PrediXcan generated a Z-score representing the association between genetically predicted DNA methylation level and ALS risk. Positive Z-scores indicate that higher genetically predicted methylation is associated with increased ALS risk, whereas negative Z-scores indicate an inverse association. The S-PrediXcan association statistic was approximated as described by Barbeira et al. [17]:

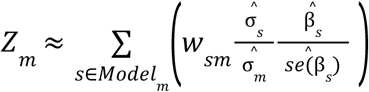

In this equation, *w_sm_* represents the weight assigned to SNP *s* in the prediction model for DNAmethylation at CpG *m*. The term β̂*_s_* and *se*(β̂*_s_*) represent the estimated effect size and corresponding standard error, respectively, for the association of SNP (s) with ALS risk in the GWAS summary statistics. The terms σ̂*_s_* and σ̂*_m_* represent the estimated standard deviations of SNP (s) and the genetically predicted methylation level at CpG site (m), respectively. Correlations among SNPs included in each prediction model were accounted for in the analysis. Statistical significance was determined using Bonferroni correction for the number of CpG sites tested, with a Bonferroni-corrected p < 0.05 considered statistically significant.

### Association analysis of genetically predicted gene expression with ALS risk

To complement the methylome-wide association analysis, we performed a transcriptome-wide association study (TWAS) to evaluate associations between genetically regulated whole-blood gene expression and ALS risk. Publicly available GTEx v8 whole-blood gene-expression prediction models were obtained from PredictDB [38]. These models use elastic net regression to predict gene expression from nearby genetic variants [16,17,30] and were applied to the same ALS GWAS summary statistics used for the methylation analyses [15].

S-PrediXcan [17] was used to estimate the association between genetically predicted expression of each gene and ALS risk. For each gene, the analysis generated a Z-score and corresponding p-value representing the direction and statistical evidence of the association. Positive Z-scores indicate that higher genetically predicted gene expression is associated with increased ALS risk, whereas negative Z-scores indicate an inverse association. Genes were evaluated when a GTEx v8 whole-blood prediction model was available and sufficient model SNPs overlapped with the ALS GWAS data. Statistical significance was determined using Bonferroni correction for the number of genes tested, with a Bonferroni-corrected p < 0.05 considered statistically significant.

## Results

### Genetically predicted blood DNA methylation identifies 25 CpGs associated with ALS risk

After applying the prediction-performance criteria from Fryett et al., 52,391 methylation prediction models from ARIES [28] and 179,965 from Understanding Society [29] were available for analysis, representing 193,315 unique CpG sites. Following quality control (R>0.1; p<0.05) and alignment with SNPs available in the ALS GWAS summary statistics, 52,088 ARIES CpGs and 179,215 Understanding Society CpGs were retained. Across both datasets, 192,378 unique CpGs were tested, including 38,925 CpGs represented in both prediction resources.

At a nominal threshold of p < 0.05, 3,741 CpGs were associated with ALS risk using the ARIES models and 13,127 using the Understanding Society models. After Bonferroni correction, 25 CpGs across eight genomic regions remained significantly associated with ALS risk (Table 1; Figures 2–3). Five were identified using ARIES models and 22 using Understanding Society models, with two CpGs (cg13958452, cg11071193) significant in both datasets. Predicted methylation was positively associated with ALS risk at 18 of the 25 sites and inversely associated at seven.

**Figure 2.**
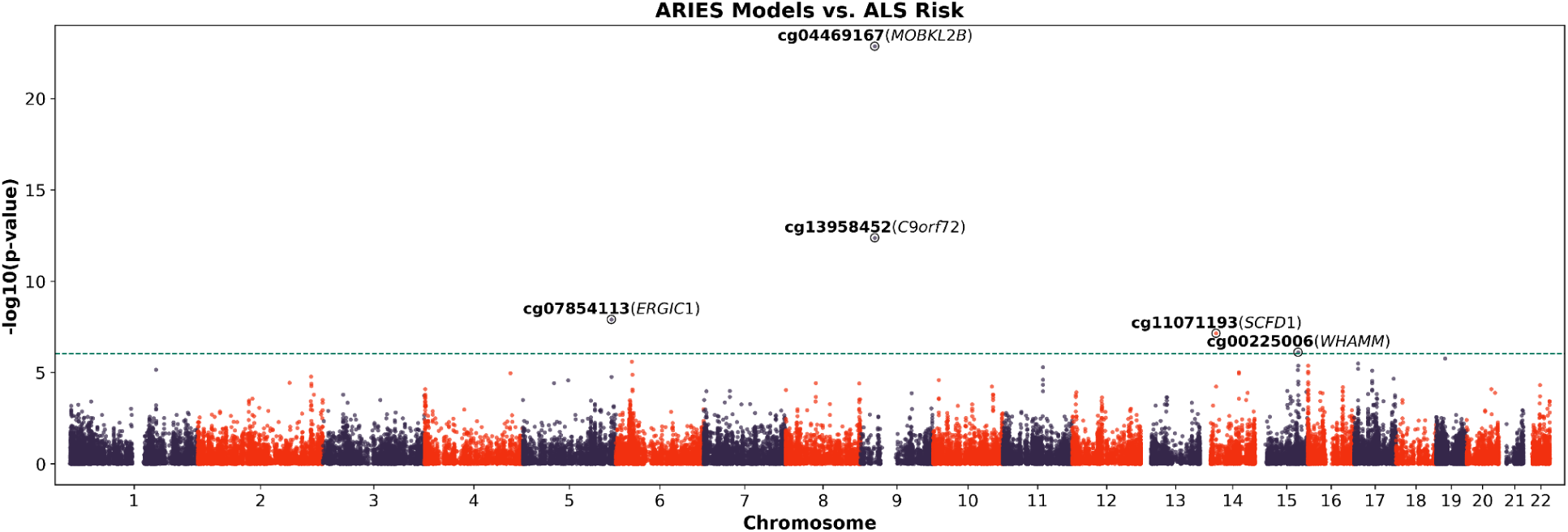
Manhattan plot of the association results from the ALS methylome-wide association study for ARIES models. The x-axis represents the genomic position of each CpG site, and the y-axis represents the -log10-transformed p-value of the association. Each dot represents the genetically predicted DNA methylation level of a specific CpG site. The red line indicates the Bonferroni-corrected significance threshold (p-value = 9.60 × 10^-7^; 0.05/52,088). CpG sites that reached the Bonferroni-corrected threshold and their nearby genes are annotated.

**Figure 3.**
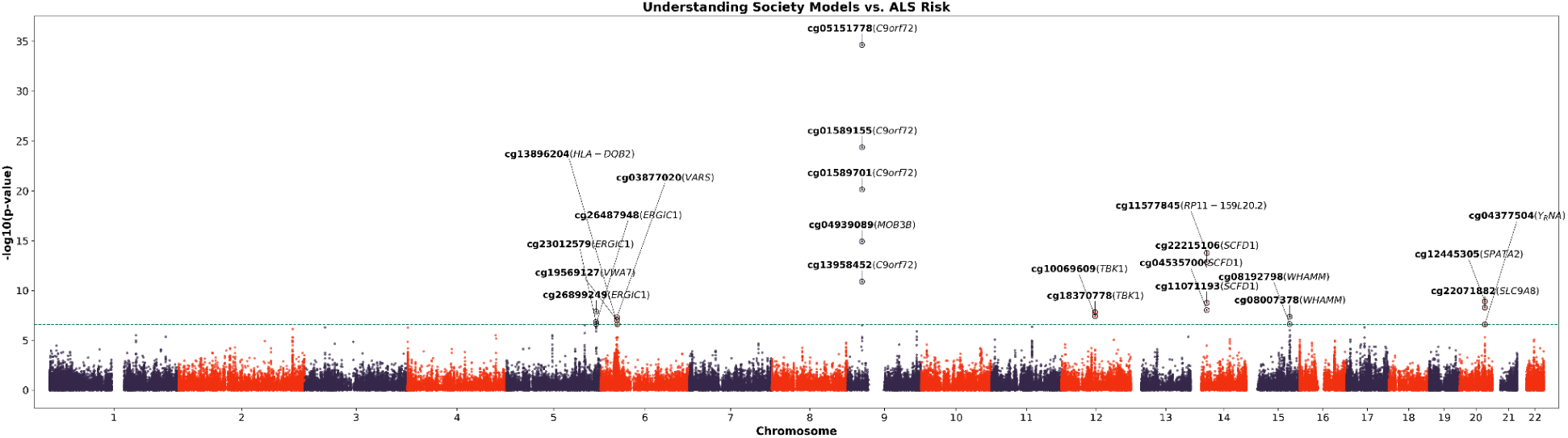
Manhattan plot of ALS methylome-wide association results using Understanding Society models. The x-axis represents the genomic position of each CpG site, and the y-axis represents the -log10-transformed p-value of the association. Each dot represents the genetically predicted DNA methylation level of a specific CpG site. The red line indicates the Bonferroni-corrected significance threshold (p-value = 2.79 × 10^-7^; 0.05/ 179,215). CpG sites that reached the Bonferroni-corrected threshold and their nearby genes are annotated.

**Table 1.** Twenty-five CpG sites significantly associated with ALS risk.

| Chr | Position | Region | CpG | Classification | Nearby Gene | ARIES (450k) |  |  | Understanding Society (EPIC) |  |  | TWAS |  |  |
| --- | --- | --- | --- | --- | --- | --- | --- | --- | --- | --- | --- | --- | --- | --- |
|  |  |  |  |  |  | Z<sup>a</sup> | P<sup>a</sup> | P<sub>Bonferroni</sub> | Z<sup>a</sup> | P<sup>a</sup> | P<sub>Bonferroni</sub> | Z<sup>a</sup> | P<sup>a</sup> | P<sub>Bonferroni</sub> |
| 5 | 172,347,685 | 5q35.1 | cg07854113 | introns | *ERGIC1* | -5.69 | 1.24E-08 | 6.45E-04 | -4.98 | 6.39E-07 | 1.15E-01 |  |  |  |
| 5 | 172,352,510 | 5q35.1 | cg26899249 | introns | *ERGIC1* |  |  |  | 5.68 | 1.33E-08 | 2.39E-03 |  |  |  |
| 5 | 172,359,545 | 5q35.1 | cg23012579 | introns | *ERGIC1* | 4.30 | 1.71E-05 | 8.92E-01 | 5.25 | 1.50E-07 | 2.70E-02 |  |  |  |
| 5 | 172,359,575 | 5q35.1 | cg26487948 | introns | *ERGIC1* |  |  |  | 5.17 | 2.33E-07 | 4.18E-02 |  |  |  |
| 6 | 31,739,837 | 6p21.33 | cg19569127 | introns | *VWA7* |  |  |  | 5.34 | 9.18E-08 | 1.65E-02 | 1.75 | 8.03E-02 | 1.00E+00 |
| 6 | 31,762,680 | 6p21.33 | cg03877020 | exons | *VARS* |  |  |  | 5.46 | 4.89E-08 | 8.76E-03 |  |  |  |
| 6 | 32,729,442 | 6p21.32 | cg13896204 | exons | *HLA-DQB2* |  |  |  | 5.16 | 2.47E-07 | 4.43E-02 | -3.19 | 1.44E-03 | 1.00E+00 |
| 9 | 27,488,839 | 9p21.2 | cg04939089 | 5'UTR | *MOB3B* |  |  |  | 8.00 | 1.21E-15 | 2.17E-10 |  |  |  |
| 9 | 27,529,907 | 9p21.2 | cg04469167 | TSS200 | *MOB3B* | 10.01 | 1.37E-23 | 7.12E-19 |  |  |  |  |  |  |
| 9 | 27,571,484 | 9p21.2 | cg13958452 | 5'UTR | *C9orf72* | 7.25 | 4.18E-13 | 2.18E-08 | 6.77 | 1.29E-11 | 2.30E-06 | 7.25 | 4.30E-13 | 3.10E-09 |
| 9 | 27,573,532 | 9p21.2 | cg01589155 | 5'UTR | *C9orf72* |  |  |  | 10.35 | 4.30E-25 | 7.71E-20 | 7.25 | 4.30E-13 | 3.10E-09 |
| 9 | 27,573,548 | 9p21.2 | cg05151778 | 5'UTR | *C9orf72* |  |  |  | 12.41 | 2.30E-35 | 4.13E-30 | 7.25 | 4.30E-13 | 3.10E-09 |
| 9 | 27,574,383 | 9p21.2 | cg01589701 | TSS1500 | *C9orf72* |  |  |  | -9.37 | 7.38E-21 | 1.32E-15 | 7.25 | 4.30E-13 | 3.10E-09 |
| 12 | 64,845,191 | 12q14.2 | cg18370778 | TSS1500 | *TBK1* |  |  |  | 5.66 | 1.52E-08 | 2.73E-03 | 5.62 | 1.94E-08 | 1.40E-04 |
| 12 | 64,852,389 | 12q14.2 | cg10069609 | introns | *TBK1* |  |  |  | -5.50 | 3.79E-08 | 6.79E-03 | 5.62 | 1.94E-08 | 1.40E-04 |
| 14 | 31,090,535 | 14q12 | cg11071193 | TSS1500 | *SCFD1* | 5.39 | 7.17E-08 | 3.74E-03 | 6.02 | 1.75E-09 | 3.14E-04 | -7.74 | 9.97E-15 | 7.20E-11 |
| 14 | 31,101,281 | 14q12 | cg22215106 | 5'UTR | *SCFD1* |  |  |  | 7.40 | 1.39E-13 | 2.50E-08 | -7.74 | 9.97E-15 | 7.20E-11 |
| 14 | 31,125,751 | 14q12 | cg04535700 | introns | *SCFD1* |  |  |  | -5.74 | 9.50E-09 | 1.70E-03 | -7.74 | 9.97E-15 | 7.20E-11 |
| 14 | 31,225,838 | 14q12 | cg11577845 | 3'UTR | *RP11-159L*<br>*20.2* |  |  |  | -7.67 | 1.72E-14 | 3.08E-09 |  |  |  |
| 15 | 83,477,872 | 15q25.2 | cg00225006 | TSS200 | *WHAMM* | 4.94 | 7.75E-07 | 4.04E-02 | 4.90 | 9.82E-07 | 1.76E-01 | -2.51 | 1.21E-02 | 1.00E+00 |
| 15 | 83,477,915 | 15q25.2 | cg08007378 | TSS200 | WHAMM | 4.60 | 4.26E-06 | 2.22E-01 | 5.48 | 4.32E-08 | 7.74E-03 | -2.51 | 1.21E-02 | 1.00E+00 |
| 15 | 83,501,835 | 15q25.2 | cg08192798 | introns | WHAMM |  |  |  | -5.17 | 2.38E-07 | 4.27E-02 | -2.51 | 1.21E-02 | 1.00E+00 |
| 20 | 48,502,892 | 20q13.13 | cg22071882 | introns | SLC9A8 |  |  |  | 5.84 | 5.26E-09 | 9.42E-04 |  |  |  |
| 20 | 48,533,123 | 20q13.13 | cg12445305 | TSS1500 | SPATA2 |  |  |  | -6.07 | 1.25E-09 | 2.24E-04 |  |  |  |
| 20 | 48,537,355 | 20q13.13 | cg04377504 | TSS200 | Y_RNA |  |  |  | 5.15 | 2.60E-07 | 4.66E-02 |  |  |  |

<sup>a</sup>Z scores and P values for association tests between genetically predicted DNA methylation or gene expression level and ALS risk.

Notably, three significant CpGs (cg00225006, cg08007378, and cg08192798) mapped to *WHAMM* at 15q25.2, were located more than 1 Mb away from any previously reported ALS GWAS risk variants. Two of them (cg00225006 and cg08007378), showed concordant positive effect directions in both the ARIES and Understanding Society models, while cg00225006 reached Bonferroni-corrected significance in both datasets.

### Genetically predicted blood gene expression identifies 11 genes associated with ALS risk

Using GTEx v8 whole-blood gene-expression prediction models [16,17,30] developed from RNA-sequencing data from 670 whole-blood samples [39], we performed TWAS analysis. We tested the genetically predicted expression level of 7,224 genes for the association with ALS risk (Figure 4). Eleven genes were significant after Bonferroni-correction (p < 6.92 × 10^-6^).

**Figure 4.**
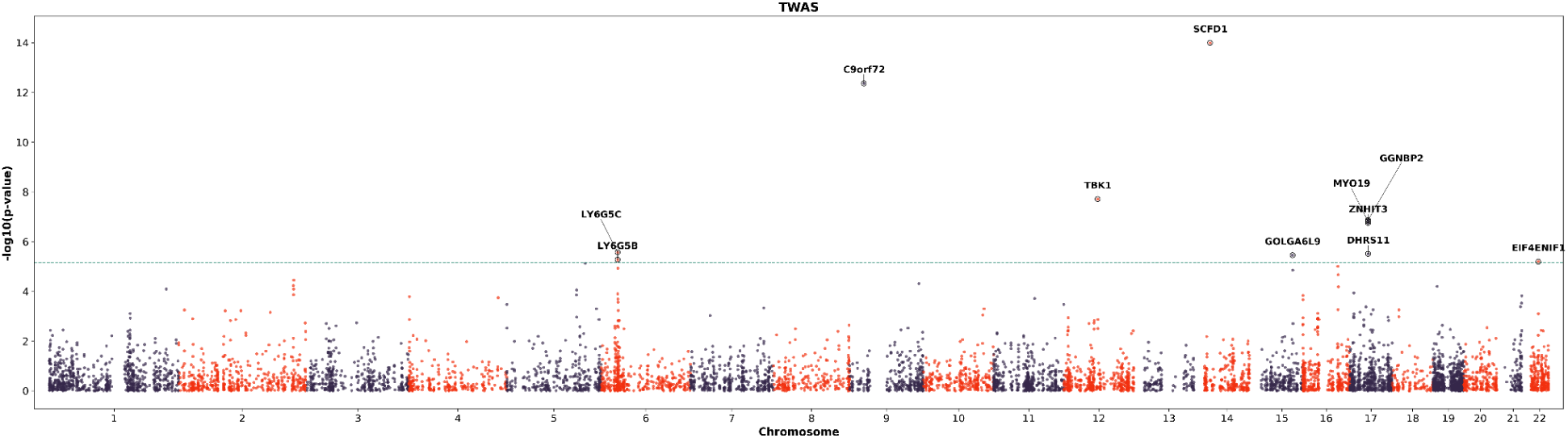
Manhattan plot of the association results from the ALS transcriptome-wide association study. The x-axis represents the genomic position of each gene, and the y-axis represents the -log10-transformed p-value of the association. Each dot represents the genetically predicted gene expression level. The red line indicates the Bonferroni-corrected significance threshold (p-value = 6.92 × 10^-6^; 0.05/ 7,224). Genes that reached the Bonferroni-corrected threshold are annotated.

Comparison of the TWAS and methylation-based association results revealed several loci represented in both analyses. At the *C9orf72* locus, cg13958452 was associated with ALS risk across both the ARIES and Understanding Society methylation models, while genetically predicted *C9orf72* expression was also associated with ALS risk in TWAS. Similarly, cg11071193, mapped near *SCFD1*, was significant across both MWAS models, while *SCFD1* was identified in the TWAS analysis.

Additional MWAS-associated CpGs mapped near *C9orf72*, *TBK1*, and *SCFD1*, genes that were also identified in the TWAS. These overlapping associations prioritize these loci for further investigation but do not establish corresponding regulatory relationships between DNA methylation and gene expression.

### MWAS-identified CpGs overlap with neurodegenerative disease GWAS loci

To evaluate whether MWAS-identified CpGs colocalized with previously reported genetic associations, we examined their proximity to genome-wide significant variants reported in the GWAS Catalog [40] for ALS, frontotemporal dementia (FTD), and Alzheimer’s disease (AD). CpGs located within 50 kb of a reported GWAS variant were considered proximal. Overall, 20 CpGs were located near 19 unique GWAS variants. Of these variants, 11 were associated with ALS, five with FTD, and four with AD, with one variant associated with both ALS and FTD. Thirteen CpGs were located within 50 kb of more than one GWAS variant, with the strongest clustering observed at the 9p21.2, 12q14.2, and 14q12 regions (Table 2).

**Table 2.** MWAS-identified CpGs located near reported ALS and other neurodegenerative disease GWAS variants.

| Disease | Chr | SNP | Region | CpG | PubmedID |
| --- | --- | --- | --- | --- | --- |
| Alzheimer's disease | 6 | rs1315875949 | 6p21.33 | cg03877020; cg19569127 | 39998322 |
| Alzheimer's disease | 6 | rs2858331 | 6p21.32 | cg13896204 | 35851147 |
| Alzheimer's disease | 6 | rs9275599 | 6p21.32 | cg13896204 | 42237039 |
| Alzheimer's disease | 20 | rs73274724 | 20q13.13 | cg04377504 | 42237039 |
| Amyotrophic lateral sclerosis | 5 | rs517339 | 5q35.1 | cg07854113; cg23012579; cg26487948; cg26899249 | 34873335 |
| Amyotrophic lateral sclerosis | 9 | rs2453555 | 9p21.2 | cg01589155; cg01589701; cg04469167; cg05151778; cg13958452 | 34873335 |
| Amyotrophic lateral sclerosis | 9 | rs2814707 | 9p21.2 | cg01589155; cg01589701; cg04469167; cg04939089; cg05151778; cg13958452 | 19734901 |
| Amyotrophic lateral sclerosis | 9 | rs3849942 | 9p21.2 | cg01589155; cg01589701; cg04469167; cg05151778; cg13958452 | 19734901; 20801718; 24256812 |
| Amyotrophic lateral sclerosis | 9 | rs3849943 | 9p21.2 | cg01589155; cg01589701; cg04469167; cg05151778; cg13958452 | 24931836; 27455348; 28931804; 29566793 |
| Amyotrophic lateral sclerosis | 9 | rs7019351 | 9p21.2 | cg04469167; cg04939089 | 25442119 |
| Amyotrophic lateral sclerosis | 12 | rs4075094 | 12q14.2 | cg10069609; cg18370778 | 34873335 |
| Amyotrophic lateral sclerosis | 12 | rs74654358 | 12q14.2 | cg10069609; cg18370778 | 29566793 |
| Amyotrophic lateral sclerosis | 14 | rs10139154 | 14q12 | cg04535700; cg22215106 | 27455348 |
| Amyotrophic lateral sclerosis | 14 | rs229194 | 14q12 | cg11071193 | 34873335 |
| Amyotrophic lateral sclerosis | 14 | rs229195 | 14q12 | cg11071193 | 34873335 |
| Frontotemporal dementia | 5 | rs517339 | 5q35.1 | cg07854113; cg23012579; cg26487948; cg26899249 | 37979250 |
| Frontotemporal dementia | 9 | rs117204439 | 9p21.2 | cg01589155; cg01589701; cg05151778; cg13958452 | 34475377 |
| Frontotemporal dementia | 9 | rs12554036 | 9p21.2 | cg04469167; cg04939089 | 37979250 |
| Frontotemporal dementia | 14 | rs229243 | 14q12 | cg04535700; cg11071193; cg22215106 | 37979250 |
| Frontotemporal dementia | 20 | rs4810992 | 20q13.13 | cg04377504; cg12445305 | 37979250 |

## Discussion

This is the first large-scale study to evaluate associations between genetically predicted blood DNA methylation and ALS risk using the PrediXcan framework. Across methylation prediction models developed from two independent reference datasets, we identified 25 CpG sites associated with ALS risk. Three of these (cg00225006, cg08007378, and cg08192798) mapped to a novel locus at 15q25.2 that has not previously been implicated in ALS by GWAS. Additional TWAS analyses integrating genetically predicted gene expression revealed concordant associations for cg13958452 and its mapped gene, *C9orf72*, further supporting the relevance of this locus to ALS susceptibility.

This framework offers a complementary approach to one of the central challenges in ALS genetics: translating GWAS signals into biologically interpretable mechanisms, particularly when associated variants lie in noncoding regions. However, a PrediXcan-based MWAS does not directly measure DNA methylation and cannot establish that methylation changes cause ALS. Rather, it tests whether the genetically regulated component of methylation is associated with disease risk. Accordingly, our results should be viewed as genetic evidence linking predicted methylation variation to ALS susceptibility and as a means of prioritizing loci for subsequent functional and biological investigation.

The first category of findings includes genes or loci with prior evidence from ALS or ALS-FTD genetic and molecular studies. Among these, two ALS-associated CpGs, cg18370778 and cg10069609, were annotated to *TBK1*. Whether through haploinsufficiency, reduced kinase activity, or disrupted protein–protein interactions, *TBK1* mutations appear to converge on a shared pathogenic pathway involving impaired autophagic clearance, proteostatic dysfunction, and accelerated neurodegeneration in ALS and FTD [41].

Three additional ALS risk-associated CpGs (cg11071193, cg22215106, and cg04535700) mapped to *SCFD1*. Previous studies have shown that *SCFD1* expression quantitative trait loci are significantly associated with increased ALS risk and that *SCFD1* expression is altered in post-mortem ALS tissue, where it correlates with disease duration [42]. *SCFD1* is a key component of a large transcriptional network linked to ALS-related pathways and genes enriched for schizophrenia risk [42], a condition known to be genetically correlated with ALS [43].

Notably, cg11577845, annotated to *RP11-159L20.2*, was associated with ALS risk. Although *RP11-159L20.2* itself has not been directly implicated in ALS, this CpG lies within the same genomic locus as the *SCFD1*-annotated CpGs, raising the possibility that these signals reflect regulatory variation within a shared ALS-relevant region.

Several other findings mapped to loci with established or emerging ALS-related evidence, with particularly strong biological support at the *C9orf72* locus. Multiple ALS risk-associated CpGs were located near *C9orf72*, including cg01589155 and cg13958452. GGGGCC repeat expansions in *C9orf72* are a major genetic cause of both ALS and FTD [44,45], with proposed pathogenic mechanisms including loss of C9orf72 function, toxic RNA foci, and accumulation of dipeptide repeat proteins [46].

Of particular interest, cg01589155 was previously associated with survival in the epigenome-wide association study by Hop et al. [47], suggesting that methylation-related variation in this locus may influence not only ALS susceptibility but also disease progression. Evidence for cg13958452 was also consistent across both the ARIES and Understanding Society methylation models, while *C9orf72* was also identified in the TWAS analysis. The consistency of these signals across methylation and gene-expression models strengthens the evidence linking molecular variation at the *C9orf72* locus to ALS susceptibility.

Additional findings mapped to loci with prior genetic or molecular support in ALS. Two ALS-associated CpGs, cg04939089 and cg04469167, were annotated to *MOB3B*, a gene located within the shared ALS-FTD risk locus at 9p21.2. In a multitrait GWAS of FTD and ALS, *MOB3B* emerged as a key candidate gene at this locus, with genetic effects contributing to susceptibility across both diseases [48]. Bayesian fine-mapping and colocalization further supported a shared causal signal at 9p21.2 (PP4 = 0.764), strengthening evidence that *MOB3B* may contribute to the common genetic architecture of ALS and FTD [48].

Support for previously implicated loci was also observed at *SLC9A8/SPATA2*, where cg22071882 mapped to *SLC9A8* and cg12445305 mapped to *SPATA2*. This finding is consistent with prior GWAS evidence implicating the *SLC9A8/SPATA2* region to ALS susceptibility *[15]*. In addition, cg13896204 was annotated to *HLA-DQB2*, an immune-related gene previously reported to be upregulated in the anterior horn of the spinal cord in sporadic ALS [49]. Together, these findings provide epigenetic support for several loci already implicated in ALS through genetic association, shared ALS–FTD susceptibility, or disease-related molecular alterations.

Several findings also pointed to pathways involved in intracellular trafficking, ER stress, and protein homeostasis. Four ALS-associated CpGs mapped to *ERGIC1*, which encodes a membrane-bound protein localized to the endoplasmic reticulum (ER)–Golgi intermediate compartment and involved in membrane trafficking and selective cargo transport between the ER and Golgi complex [50]. Disruption of ER–Golgi transport has been reported as a shared pathogenic mechanism across SOD1-, TDP-43-, and FUS-associated ALS [51], while ER stress has also been implicated more broadly in ALS pathogenesis [52].

A separate signal, cg03877020, mapped to *VARS* and was positively associated with ALS risk. VARS1 encodes valine-tRNA ligase and has been identified among translational proteins showing increased affinity for cytoplasmic TDP-43 [53]. Given that nuclear depletion and cytoplasmic accumulation of TDP-43 are central pathological features of ALS, this observation raises a potential connection between VARS1 and TDP-43-associated cellular dysfunction. However, the available evidence derives from protein-interaction studies in cell lines and does not establish that VARS1 co-localizes with TDP-43 aggregates or contributes directly to ALS pathogenesis.

Additional candidate loci were supported by prior molecular evidence in ALS. The ALS risk-associated CpG cg19569127 mapped to *VWA7*, a gene previously identified among 44 transcripts that were significantly differentially expressed across two independent ALS post-mortem motor cortex RNA-seq datasets, KCL BrainBank and TargetALS [54]. *VWA7* was downregulated in ALS relative to controls in both cohorts, with log₂ fold changes of −0.24 and −0.47, respectively (adjusted p = 0.0284 in both datasets), and was included in an extracellular matrix-related transcriptional signature shared across the two cohorts [54].

Three additional CpGs (cg00225006, cg08007378, and cg08192798) mapped to *WHAMM* within 15q25.2, a region not previously implicated in ALS through GWAS. Although this represents a potentially novel ALS-associated locus, *WHAMM* has previously been proposed as a cell-type–specific candidate gene with possible relevance to ALS pathogenesis [55]. Another ALS-associated signal, cg04377504, mapped to Y RNA. Supporting the potential relevance of this finding, Y RNA abundance has been reported to differ significantly between individuals with ALS and controls in both plasma and serum [56]. These observations highlight additional loci and molecular features that warrant further investigation but currently have more limited evidence linking them directly to ALS biology.

The proximity of several MWAS-identified CpGs to previously reported ALS and FTD GWAS variants indicates that a subset of the methylation signals falls within regions already implicated in neurodegenerative disease. However, genomic proximity alone does not establish that these variants regulate the corresponding CpGs or that the associations reflect the same underlying causal signal. Further analyses incorporating mQTL data, linkage disequilibrium structure, and colocalization will be needed to determine whether the genetic and methylation associations share common regulatory mechanisms.

Several limitations should be considered when interpreting these findings. First, the methylation prediction models were derived from blood rather than brain or spinal cord tissue. Although blood-based models may capture systemic components of ALS biology, they may not reflect genetically regulated methylation patterns in motor neurons, glial cells, or other central nervous system cell types. Development and application of tissue- and cell-type–specific prediction models will therefore be important for determining whether the associations identified here are also relevant within disease-affected tissues. Second, the direction of an MWAS association should not be interpreted as indicating an activating or repressive regulatory effect. The functional consequences of DNA methylation depend on the genomic location and regulatory context of the CpG site. Consequently, the directions of the MWAS and TWAS associations cannot be assumed to represent directly comparable regulatory effects. Finally, the analyses were restricted to individuals of European ancestry, which may limit the generalizability of the findings. Future studies incorporating larger and more ancestrally diverse populations will be needed to assess the robustness and transferability of these associations.

In summary, this study provides a systematic evaluation of genetically predicted blood DNA methylation in relation to ALS risk. By integrating methylation prediction models with large-scale ALS genetic data, we identified signals spanning established ALS genes and loci, regions linked to ALS-relevant biological pathways, and less characterized candidate loci that may warrant further investigation. These results illustrate how PrediXcan-based MWAS can complement conventional GWAS by helping prioritize genetically regulated molecular signals within or near associated regions, particularly where the functional consequences of underlying genetic variation remain unclear. Importantly, these associations should not be interpreted as evidence that altered DNA methylation itself causes ALS, but rather as a framework for prioritizing loci and molecular mechanisms for subsequent functional validation.

## Conclusions

In this first large-scale PrediXcan study of genetically predicted blood DNA methylation and ALS risk, we identified 25 CpG sites associated with ALS, including three at the 15q25.2 locus, a region not previously implicated by ALS GWAS. Complementary TWAS analyses further revealed convergent regulatory signals at *C9orf72*.

The broader impact of these findings is that they add an epigenetic regulatory dimension to the interpretation of ALS genetic risk. Rather than identifying additional risk variants alone, this approach helps prioritize specific CpG sites, genes, and genomic regions through which inherited genetic variation may influence disease susceptibility. In doing so, PrediXcan-based MWAS can help refine broad GWAS loci into more biologically interpretable candidates, highlight regulatory mechanisms that may otherwise be missed by variant-level analyses, and identify targets for functional validation. This is particularly relevant in ALS, where many associated variants lie in noncoding regions and their biological consequences remain poorly understood. By linking genetic variation to predicted methylation and integrating these signals with gene-expression analyses, our study provides a complementary strategy for moving from genetic association toward mechanistic understanding of ALS susceptibility.

## Data Availability

All data produced in the present study are available upon reasonable request to the authors.

https://www.ebi.ac.uk/gwas/studies/GCST90027164

https://www.bristol.ac.uk/alspac/researchers/access/

https://www.understandingsociety.ac.uk/documentation/health-assessment/accessing-data/

https://www.staff.ncl.ac.uk/heather.cordell/MethPaper.html

https://predictdb.org

## List of abbreviations

ALS: Amyotrophic lateral sclerosis
ARIES: Accessible Resource for Integrated Epigenomic Studies
CpG: Cytosine–phosphate–guanine
ER: Endoplasmic reticulum
FTD: Frontotemporal dementia
GTEx: Genotype-Tissue Expression
GWAS: Genome-wide association study
MWAS: Methylome-wide association study
RNA-seq: RNA sequencing
SNP: Single-nucleotide polymorphism
TWAS: Transcriptome-wide association study

## Declarations

## Ethics approval and consent to participate

Not applicable. This study used publicly available, deidentified summary-level data and did not involve the enrollment of participants or analysis of individual-level data by the authors. Ethics approval and informed consent for the original studies are described in the respective source publications.

## Consent for publication

Not applicable.

## Availability of data and materials

The data that support this study are available from the corresponding author upon reasonable request. Publicly available software and packages were used throughout this study according to each developer’s instructions. Custom scripts generated for the study will be available upon request.

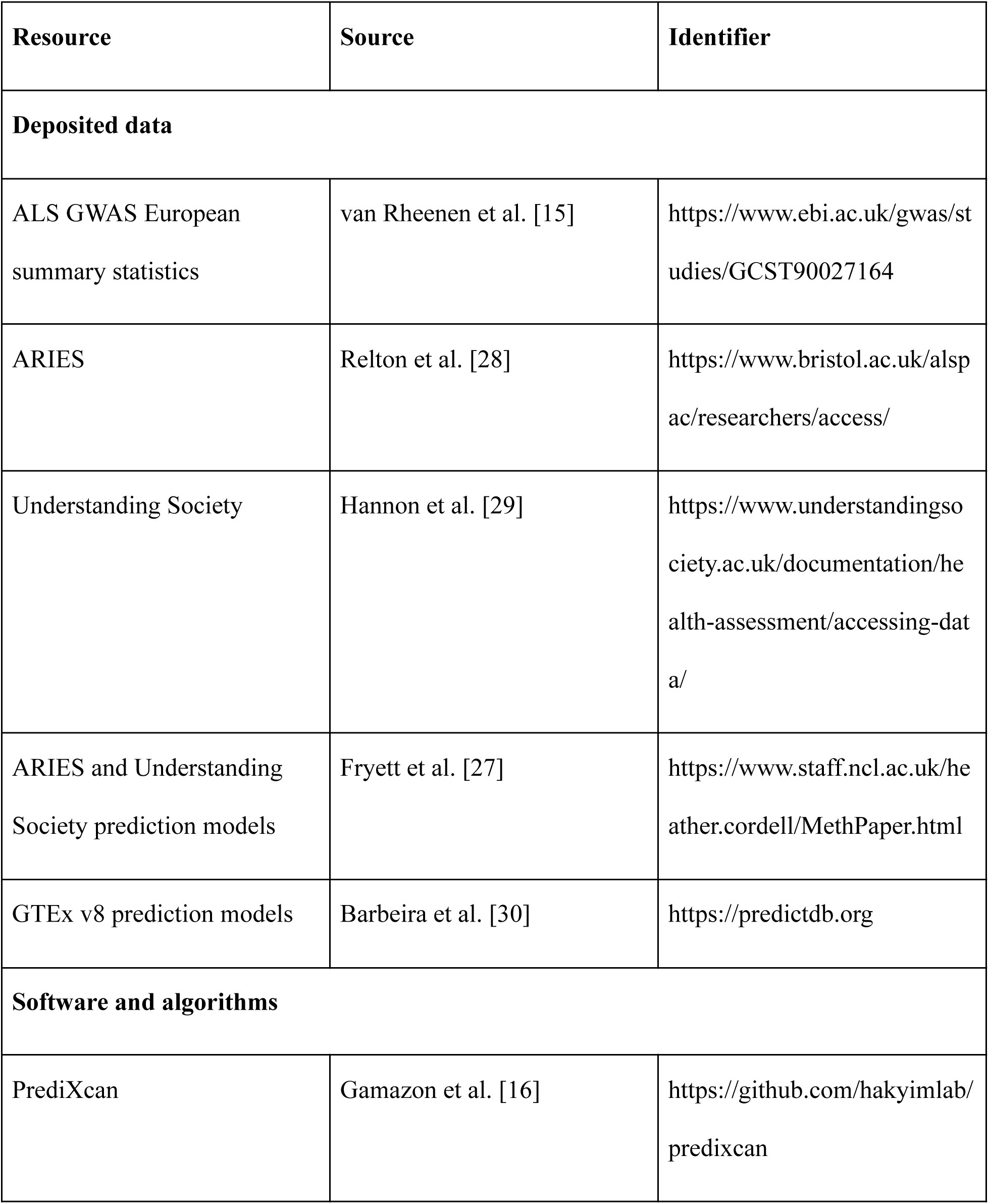
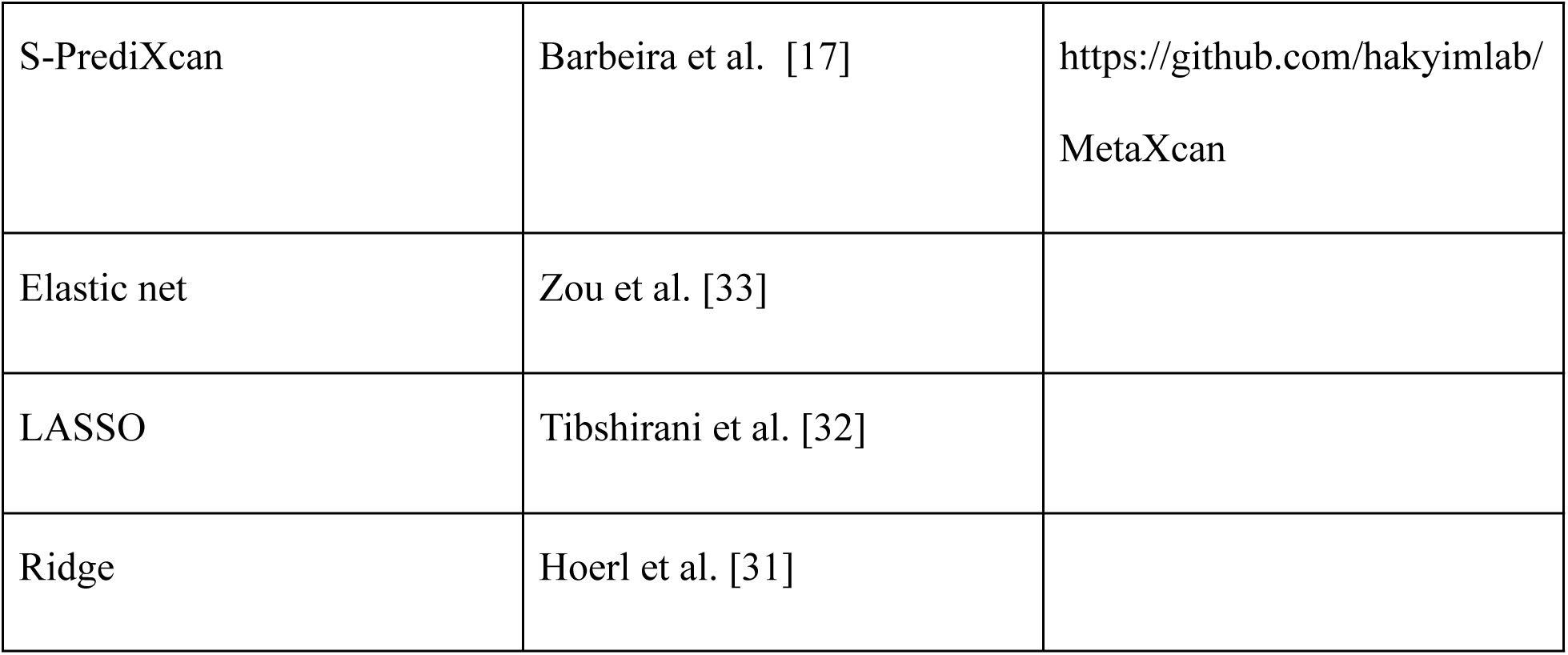

## Authors’ contributions

Tianying Zhao performed all analyses, interpreted the results, and drafted the manuscript. Mariah Hoffman, Gang Wu, and Veronique Belzil provided supervision, scientific guidance, and critical revision of the manuscript.

## Acknowledgements

The authors gratefully acknowledge the investigators and participants whose contributions made the publicly available datasets used in this study possible.

